# Neighbourhood deprivation across eight decades and late-life cognitive function in the Lothian Birth Cohort 1936: A life-course study

**DOI:** 10.1101/2022.08.16.22278836

**Authors:** Gergő Baranyi, Federica Conte, Ian J. Deary, Niamh Shortt, Catharine Ward Thompson, Simon R. Cox, Jamie Pearce

## Abstract

**Introduction:** Although neighbourhood may predict late-life cognitive function, studies mostly relies on measurements at a single time point, with few investigations applying a life-course approach. Further, it is unclear whether the associations between neighbourhood and cognitive tests scores relate to specific cognitive domains or general ability. This study explored how neighbourhood deprivation across eight decades contributes to late-life cognitive function.

**Methods:** Data were drawn from the Lothian Birth Cohort 1936 (n=1091) with cognitive function measured through 10 tests at ages 70, 73, 76, 79 and 82. Participants’ residential history was gathered with ‘lifegrid’ questionnaires and linked to neighbourhood deprivation in childhood, young adulthood, and mid-to-late adulthood. Associations were tested with latent growth curve models for levels and slopes of general (*g*) and domain-specific abilities (visuospatial ability, memory, and processing speed), life-course associations were explored with path analysis.

**Results:** Higher mid-to-late adulthood neighbourhood deprivation was associated with lower age 70 levels (*β*=-0.113, 95%CI: −0.205, −0.021) and faster decline of *g* over 12 years (*β*=-0.160, 95%CI: −0.290, −0.031). Initially-apparent findings with domain-specific cognitive functions (e.g. processing speed) were due to their shared variance with *g*. Path analyses suggested that childhood neighbourhood disadvantage is indirectly linked to late-life cognitive function through education and selective residential mobility.

**Conclusions:** To our knowledge, we provide the most comprehensive assessment of the relationship between life-course neighbourhood deprivation and cognitive ageing. Living in advantaged areas in mid-to-late adulthood may contribute to better cognitive function and slower decline, whereas advantaged childhood neighbourhood environment likely affects functioning through cognitive reserves.

## INTRODUCTION

Mean lifespan changes in cognitive functioning have been observed in healthy non-clinical older adults, with fluid abilities such as reasoning, memory, and processing speed peaking in early adulthood and declining throughout the second part of life. Crystallised abilities, relying on previously-acquired knowledge and skills, tend to decline in mean levels well after the 60s.^1-3^ Non-pathological cognitive ageing is much more prevalent than dementia, it affects everyday functioning, quality of life and independent living, and it can herald illness, dementia, and death.^4-6^ Identifying and targeting modifiable life-course risk factors of non-pathological cognitive ageing may slow cognitive decline, reduce personal and societal burden, and contribute to healthy population ageing.

Several neighbourhood features have been proposed as modifiable determinants of late-life cognitive function, with the majority of studies focussing on area-disadvantage.^7-10^ However, reviews consistently report the lack of repeatedly-measured exposures,^8-10^ restricting understanding of when in the life course place-based factors might shape outcomes;^9^ neighbourhoods change over time, people selectively move between places, and exposures might have long-lasting impact given their timing during development.^11^ The life-course approach might help to understand long-term associations, but few investigations have assessed possible sensitive periods in the context of environment and cognitive functioning^11-13^ or dementia,^14^ and, to our knowledge, no study has focussed on neighbourhood disadvantage over the life course and non-pathological cognitive ageing. Moreover, studies usually investigate direct associations after adjustments for confounders,^9^ but it is plausible that neighbourhoods, especially during the first half of life, indirectly contribute to late-life cognition via education or socioeconomic position.

Existing literature is further limited by brief and relatively insensitive cognitive measurements (e.g. MMSE);^9,10^ whereas comprehensive assessments across multiple domains of functioning are rare, especially in longitudinal setting. Research also needs to consider that cognitive domains are positively intercorrelated, suggesting an underlying general ability factor which accounts for much of the variation between cognitive tests^2^ and their change over time.^15^ Overcoming these concerns requires robust measurements of specific cognitive domains utilising multiple repeatedly administered tests, and greater understanding of whether associations are domain-specific, or pervasive across domains linking findings to general cognitive ability (*g*).

This study explored the life-course relationship between neighbourhood deprivation and non-pathological late-life cognitive function, in a Scottish birth cohort with area- and individual-level data across eight decades. First, we quantified associations between life-course neighbourhood deprivation and the levels of *g* and domain-specific cognitive function (visuospatial ability, memory, and processing speed) at age 70, and their trajectories over five testing waves between age 70 and 82. Second, we tested whether there were unique (non-general) associations with specific cognitive domains, by separating the cognitive tests’ shared (general) variance from residual variance attributable to single domains. Last, we explored life-course pathways between area- and individual-level socioeconomic position, and cognitive function.

## METHODS

### Study participants

We obtained data from the Lothian Birth Cohort 1936 (LBC1936), a longitudinal study of community-dwelling, relatively healthy older Scottish adults, established to study individual differences in cognitive ageing.^16^ The sample was recruited with the aim of re-examining some participants of the Scottish Mental Survey 1947 (SMS1947), a nation-wide school-based cognitive ability test of 1936-born Scottish schoolchildren, carried out on June 4, 1947.^16^ Potentially surviving participants of SMS1947 living in the Lothian region of Scotland (including Edinburgh) were retraced and contacted by the Lothian Health Board.^17^ The baseline wave took place between 2004 and 2007, and included 1091 men and women with the average age of 70 years. Since then, participants have been re-examined at age 73 (2007-2010; n=866), 76 (2011-2013; n=697), 79 (2014-2017; n=550), and 82 (2017-2020; n=431).^17^

### Life-course neighbourhood social deprivation

At the mean age of 78 (2014), a ‘lifegrid’ questionnaire was administered to surviving LBC1936 participants collecting information on residential history for each decade from birth to date of completion.^17^ Recall was assisted by ‘flashbulb’ memory prompts (e.g. 9/11 attacks in New York), and by giving participants the option to write down key personal events.^12^ Out of 704 contacted cohort members, 593 provided usable life grid data; 7423 addresses were geocoded with automatic geocoders and historical building databases.^12,18^

Decade-specific neighbourhood social deprivation (NSD) scores were constructed for the City of Edinburgh (see details elsewhere).^18^ Briefly, 1941, 1951, 1961 and 1971 NSD was captured using a historical index of multiple deprivation (i.e. population density, overcrowding, infant mortality, tenure, and amenities);^18^ 1981, 1991, 2001 and 2011 NSD using the Carstairs Index of Deprivation (i.e. male unemployment, overcrowding, car ownership, and social class).^19^ Data were derived from historical records, and aggregated into a common spatial resolution (1961 census wards; n=23) to support missing data imputation.^18^ Decade-specific NSD values were transformed into *z*-scores to ensure comparability.^18^

We linked NSD scores to participants’ residential history using time bands of 10 years (e.g. 1941 score to 1936-1945 addresses). There was very high correlation between individual scores closer in time; therefore, we computed average exposure in childhood (1936-1955; age 0-19), young adulthood (1956-1975; age 20-39), and mid-to-late adulthood (1976-2014; age 40-78) (Supplementary Figure 1).

### Cognitive abilities

We utilised ten cognitive tests, administered individually across all five study waves at the same location, using the same instruments, and the same instructions.^16^ Following previous work on their correlational structure,^20^ tests were grouped into three domains:

*Visuospatial ability* was captured with the Block Design and Matrix Reasoning tests from the Wechsler Adult Intelligence Scale, 3rd UK Edition (WAIS-III^UK^)^21^ and the Spatial Span test (average of forwards and backwards) from Wechsler Memory Scale, 3rd UK Edition (WMS-III^UK^).^22^

*Memory* was measured with the Logical Memory and the Verbal Paired Associates tests from WMS-III^UK^ (total score of immediate and delayed, in each case), and with the Backward Digit Span test from WAIS-III^UK^.

*Processing speed* included the Digit Symbol Substitution and the Symbol Search tests from the WAIS-III^UK^ and two experimental tasks: Four-Choice Reaction Time^23^ and Inspection Time.^24^ Detailed information on these tests can be found elsewhere.^16^

### Covariates

Three sets of covariates were considered (Supplementary Figure 2). The first included age, sex, parental occupational social class (OSC) (professional-managerial [I/II], skilled, partly skilled and unskilled [III/IV/V])^25^, and apolipoprotein E (*APOE*) ε4 allele status (ε4 carriers, not ε4 carriers); potential confounders for all NSD-cognitive function associations. The second set included childhood IQ (measured as part of the SMS1947 at age 11 with the Moray House test No.12),^16^ years spent in (full-time) education, and adult OSC (I/II, III/IV/V);^25^ these are covariates that might mediate the association of earlier exposures, but also might confound later exposure. The final set of covariates comprised variables capturing health-related conditions measured at LBC1936 baseline (i.e. age 70): smoking status (current smoker, ex-smoker, never smoked), body mass index (BMI), and history (yes, no) of self-reported medical diagnoses (cardiovascular disease; diabetes; hypertension; and stroke). As they were concurrently measured with the last NSD epoch (age 40-78) and we could not ascertain that they were true confounders, as opposed to being mediators, we only included them in the sensitivity analysis.

### Statistical analysis

Data were analysed with latent growth curve modelling within a structural equation modelling framework. We applied the hierarchical ‘factor-of-curves’ approach^26^ (see example in Figure 1A), which has been previously used with the LBC1936 cohort.^27,28^ Levels (i.e. intercepts at age 70) and slopes (i.e. trajectories between age 70 and 82) of individual cognitive tests’ scores were calculated. Linear slopes were modelled by setting the path from the slope to the baseline test score to zero, and using the average time lag between follow-up waves as path weights (i.e. wave1-2: 2.98 years; wave1-3: 6.75 years; wave1-4: 9.82 years; wave1-5: 12.54 years). Test levels and slopes loaded onto levels and slopes of higher-order latent domains (visuospatial ability, processing speed, and memory), latent domains loaded onto a higher-order *g* capturing their common variance. Models were fitted with correlations allowed between levels and slopes, for both measured and latent variables. Time-variant age (standardized and centred within-wave) was specified at the measurement level to account for participants’ modest age differences at attendance of each wave; time-invariant covariates and exposures were fitted at the latent domain level as linear regressions. In the main analysis, two nested models were run for each domain separately: Model 1 adjusted for the first (i.e. age, sex, parental OSC, *APOE* ε4 allele status), and Model 2 adjusted for an additional set of covariates (i.e. childhood IQ, years spent in education, adult OSC).

**Figure 1:**
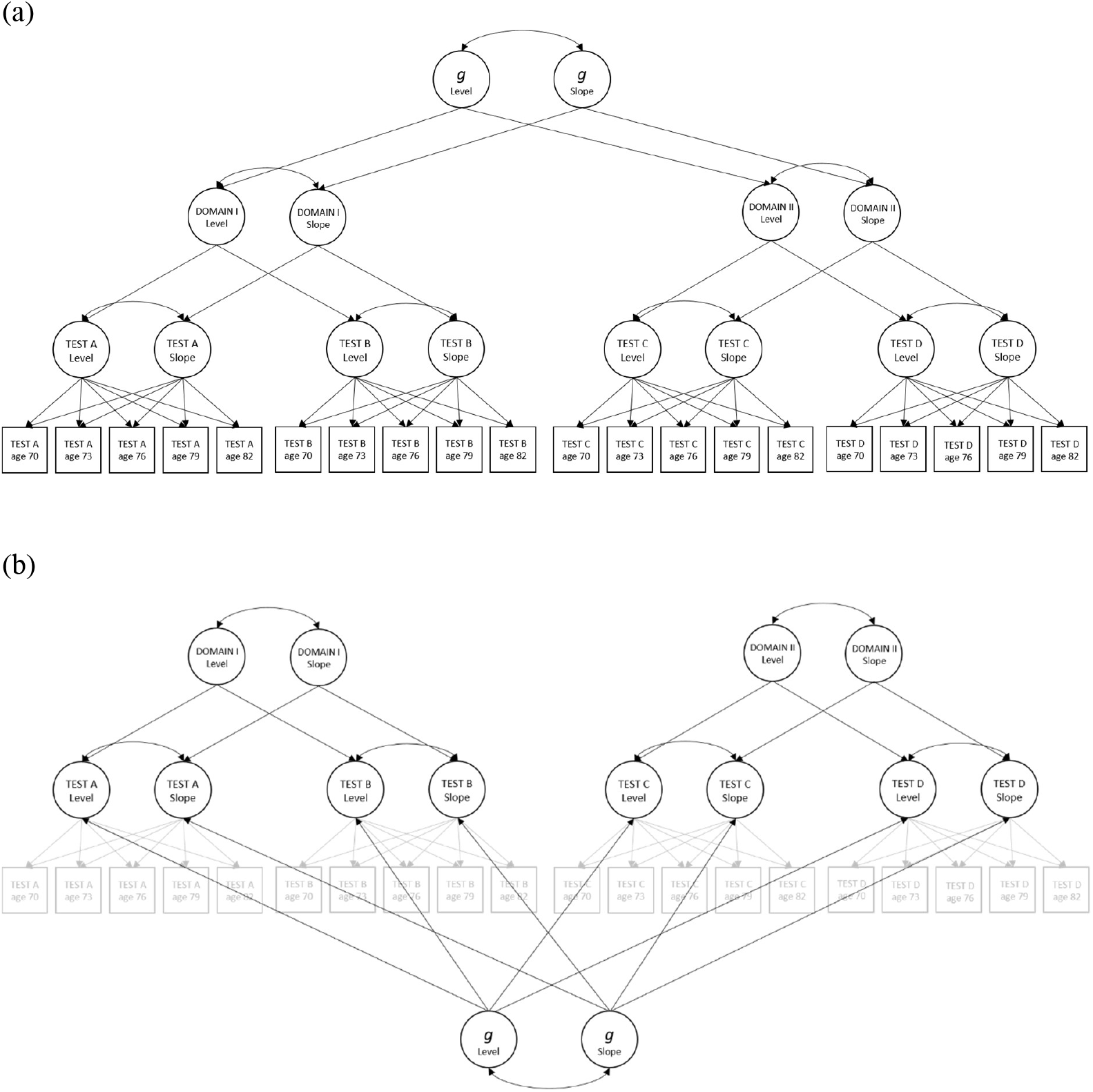
Simplified diagrams of the (a) ‘factor-of-curves’ model and the (b) bifactor model of cognitive ability. Diagram **(a)** represents a simplified ‘factor-of-curves’ model. A growth curve for each cognitive test was estimated with latent levels and slopes; loadings on the test slopes were set to 0, 2.98, 6.75, 9.82 and 12.54 to represent the average time passed between wave 1 and follow-up assessments. Test levels and slopes loaded onto levels and slopes of domain-specific cognitive abilities, which loaded onto levels and slopes of general ability (*g*). Models were run separately for each domain (i.e. visuospatial ability, verbal memory, processing speed) and for *g*. Diagram **(b)** represents a simplified bifactor model. Latent growth curves were constructed for cognitive tests similarly to the ‘factor-of-curves’ model. The variance of latent test levels and slopes were partitioned into variance contributing to specific domains and contributing to *g*; model was run simultaneously for domains and *g*. Time-variant age (i.e. standardized and centred) was included at the measurement level; time-invariant covariates and exposures of interest at the levels and slopes of latent hierarchical domains as regression equations. Variables in squares are measured, variables in circles are latent; double headed arrows represent correlations.

A previous LBC1936 study with five waves of cognitive test scores found that a latent *g* accounts for 43% of the variability in the levels and 71% of the variability in the slopes of individual test scores.^29^ Given these intercorrelations, we further assessed to what extent associations between NSD and cognitive domains were simply reflective of a pervasive association across all cognitive domains (i.e., *g*), or whether there were any domain-specific cognitive associations beyond that. To do so, we ran a longitudinal bifactor model which builds on the foundation of the ‘factor-of-curves’ approach (see example in Figure 1B) but partitions the test variance loading onto latent levels and slopes of *g* from those uniquely contributing to specific domains.^28^ In the bifactor model, the associations of NSD with domain-specific and general abilities were simultaneously tested. Before including time-invariant covariates on the latent variables, we fixed loadings and covariances to values estimated in the measurement model (i.e. model without time-invariant covariates) to ensure model convergence.

We fitted a path model with associations between exposures, outcomes, and potential mediators (i.e., childhood IQ, education, adult OSC) modelled in their life-course order, and controlled for Model 1 confounders. Levels and slopes of *g* for each of the participants were predicted and extracted from the ‘factor-of-curves’ measurement model. Due to temporal overlap in measurement, we specified the link between childhood NSD and childhood IQ as correlation.

Three sets of analyses assessed the robustness of the ‘factor-of-curves’ models. First, we additionally included the third set of covariates capturing health-related conditions. Second, to capture healthy cognitive ageing (and to reduce the likelihood of recall bias on residential history), we excluded individuals with signs of cognitive impairment, defined as either reporting the diagnosis of dementia or scoring <24 on the MMSE in any of the study waves. Third, as an alternative operationalisation of NSD, we calculated residualised change between epochs (i.e. young adulthood residualised on childhood, mid-to-late adulthood on young adulthood), and reran the main models.

Models were run with full information maximum likelihood estimation to consider all available data, and effect estimates were reported as standardized coefficients (*β*s) with their 95% confidence intervals (CI). We assessed model fit with four absolute fit indices: Comparative Fit Index (>0.95 considered as acceptable), Tucker-Lewis Index (>0.95), Root Mean Square Error of Approximation (<0.06), and Standardized Root Mean Square Residual (<0.08).^30^ To reduce type 1 errors, we provided false discovery rate (FDR) adjusted p-values.^31^ Analyses were implemented using *lavaan* package^32^ in R version 4.1.2.^33^

## RESULTS

### Descriptive statistics

Out of 1091 individuals participating in the baseline wave, 50.23% were male, 27.08% had lower parental OSC, and 29.77% were *APOE* ε4 carriers. As residential history was first collected after wave 3, and deprivation scores could only be linked to those residing in Edinburgh, the number of individuals with missing information on childhood, young adulthood and mid-to-late adulthood NSD was substantial (Table 1). Raw score means and standard deviations of the longitudinal cognitive tests are presented in Supplementary Table 1 for the whole sample; in Supplementary Table 2 for completers only (i.e. individuals present at all 5 waves). Cognitive test scores between age 70 and 82 declined across all measured tests (as reported previously^29,34^).

**Table 1:** Descriptive statistics

| | Variable | N total | Mean ± SD / N (%) |
| --- | --- | --- | --- |
| Exposure | Neighbourhood social deprivation, mean ± SD | | |
| | Childhood | 400 | 0.387 ± 3.28 |
| | Young adulthood | 482 | -0.863 ± 2.73 |
| | Mid-to-late adulthood | 494 | -2.203 ± 2.82 |
| Covariates I | Age, mean ± SD | 1091 | 69.53 ± 0.83 |
|  | Sex, n (%) | 1091 |  |
|  | Male |  | 548 (50.23%) |
|  | Female |  | 543 (49.77%) |
|  | Parental occupational social class, n (%) | 960 |  |
|  | I and II |  | 260 (27.08%) |
|  | III, IV and V |  | 700 (72.92%) |
| | *APOE* ε4 allele status, n (%) | 1028 | |
| | ε4 carriers | | 306 (29.77%) |
| | Not ε4 carriers | | 722 (70.23%) |
| Covariates II | Childhood IQ, mean ± SD | 1028 | 100.00 ± 15.00) |
| | Years spent in education, mean ± SD | 1091 | 10.74 ± 1.13) |
|  | Adult occupational social class, n (%) | 1070 |  |
|  | I and II |  | 592 (55.33%) |
|  | III, IV and V |  | 478 (44.67%) |
| Covariates III | Smoking status at baseline, n (%) | 1091 |  |
|  | Current smoker |  | 125 (11.46%) |
|  | Ex-smoker |  | 465 (42.62%) |
|  | Never smoked |  | 501 (45.92%) |
| | BMI at baseline, mean ± SD | 1089 | 27.78 ± 4.36 |
|  | History of cardiovascular diseases, n (%) | 1091 |  |
|  | Yes |  | 268 (24.56%) |
|  | No |  | 823 (75.44%) |
|  | History of diabetes, n (%) | 1091 |  |
|  | Yes |  | 91 (8.34%) |
|  | No |  | 1000 (91.65%) |
|  | History of hypertension, n (%) | 1091 |  |
|  | Yes |  | 433 (39.69%) |
|  | No |  | 658 (60.31%) |
|  | History of stroke, n (%) | 1091 |  |
|  | Yes |  | 54 (4.94%) |
|  | No |  | 1037 (95.05%) |

\*Model adjusting for Covariates III is presented in the sensitivity analysis.

**Table 2:**
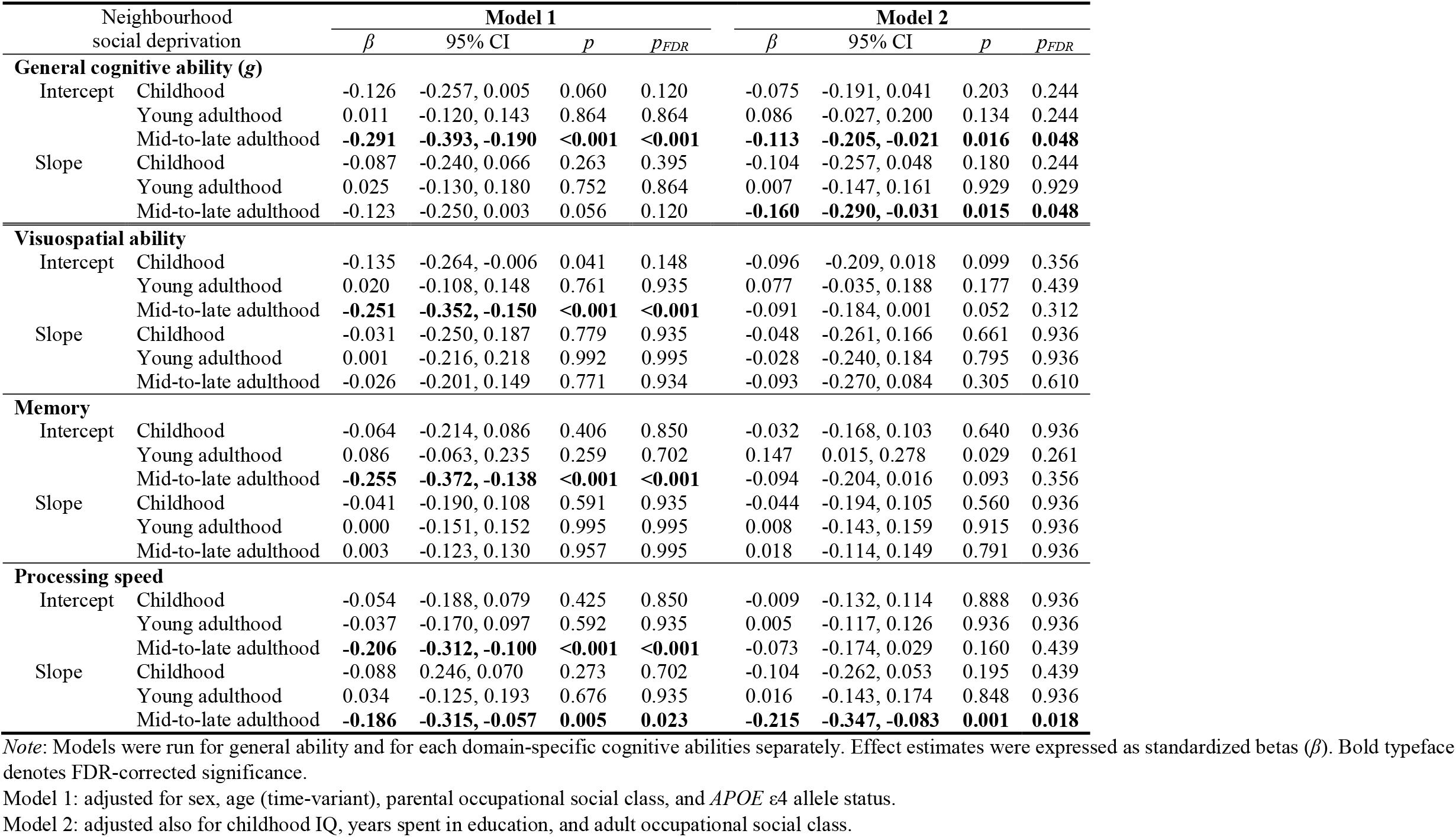
‘Factor-of-curves’ models for the association between neighbourhood social deprivation and cognitive function

### Is life-course neighbourhood social deprivation associated with cognitive function*?*

Figure 2—which is used for description and not for analysis—depicts the unadjusted associations between intercepts and slopes of *g* and NSD in three epochs. A linear relationship was observed for childhood NSD and intercept and slope of *g*, whereas in mid-to-late adulthood differences in cognition were more pronounced between low versus moderate/high levels of deprivation. In young adulthood, differences were less apparent.

**Figure 2:**
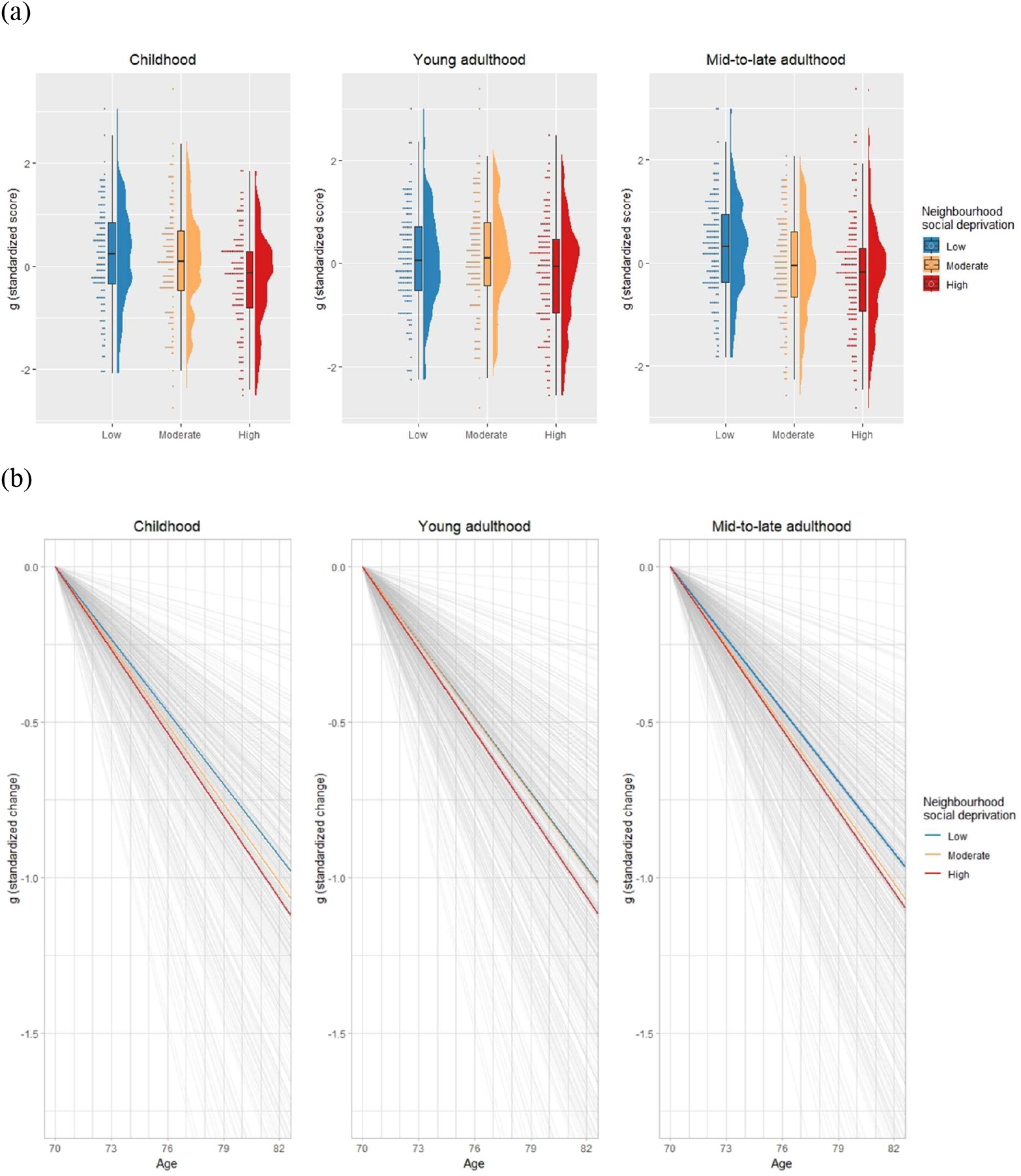
Average (a) intercepts and (b) slopes of general cognitive ability (*g*) by low, moderate and high levels of neighbourhood social deprivation in childhood, young adulthood and mid-to-late adulthood. Slopes and intercepts of *g* were extracted from the ‘factor-of-curves’ measurement model without adjustment of time-invariant covariates. For this figure, intercepts were centred and standardized, slopes are expressed as standardized change (i.e. raw slope values divided by the standard deviation of the raw intercept values). Continues measures were split into three equal groups representing low, moderate and high deprivation. Estimates are based on *n*=400 for childhood, *n*=482 for young adulthood, and on *n*=494 for mid-to-late adulthood deprivation.

After adjusting for sex, age, parental OSC and *APOE* ε4 allele status (Model 1), higher mid-to-late adulthood NSD was associated with lower level of *g* (*β*=-0.291; 95%CI: −0.393, −0.190), visuospatial ability (*β*=-0.251; 95%CI: −0.352, −0.150), memory (*β*=-0.255; 95%CI: −0.372, - 0.138) and processing speed (*β*=-0.206; 95%CI: −0.312, −0.100), and with steeper declines in processing speed (*β*=-0.186; 95%CI: −0.315, −0.057) (Table 2). On further adjustment for childhood IQ, education, and adult OSC (Model 2), the associations between mid-to-late adulthood neighbourhood deprivation and the intercept (*β*=-0.113; 95%CI: −0.205, −0.021) and slope of *g* (*β*=-0.160; 95%CI: −0.290, −0.031) as well as the slope of processing speed (*β*=-0.215; 95%CI: −0.347, −0.083) passed FDR correction for multiple comparison (Table 2); the magnitude of the associations was comparable or larger than for indicators of individual socioeconomic status (Supplementary Table 3). Models showed good fit to the data, with a few exceptions of marginally poorer fits (all CFI>0.933, all TLI>0.930) than stated cutoffs^30^ (Supplementary Table 4).

### Does general cognitive function account for the associations of domain-specific abilities?

In the longitudinal bifactor model we removed the test variance of *g* from domain-specific abilities, to test whether domain-specific findings from the ‘factor-of-curves’ models were reflecting pervasive, cross-domain, cognitive associations or whether there were additional, unique cognitive domain associations with NSD. Models suggested that findings were likely due to their shared variance with *g*: only the associations for *g* remained significant in the bifactor model before FDR correction (with similar direction and comparable magnitude as in the main ‘factor-of-curves models) and the association between mid-to-late adulthood NSD and processing speed in the fully adjusted model dropped from *β*=-0.215 to *β*=0.001 (Supplementary Table 5).

### How does life-course neighbourhood deprivation contribute to late-life cognition across the lifespan?

We fitted a path model to explore direct or indirect associations between life-course neighbourhood deprivation and late-life cognitive function. After inspecting the correlational structure of variables, we specified all potential pathways between exposure, mediators, and intercept and slope of *g*. Childhood NSD was correlated with childhood IQ (*β*=-0.151). Although there was no direct effect, higher childhood NSD had a downstream association with *g* through two pathways. First, it was associated with shorter time spent in education (*β*=-0.143) predicting *g* intercept (*β*=0.111). Second, it contributed to young adulthood NSD (*β*=0.540), which, in turn, to mid-to-late adulthood NSD (*β*=0.326); and—as shown in the ‘factor-of-curves’ model—mid-to-late adulthood NSD was associated with the intercept (*β*=-0.095) and slope of *g* (*β*=-0.115) (Figure 3; Supplementary Table 6).

**Figure 3:**
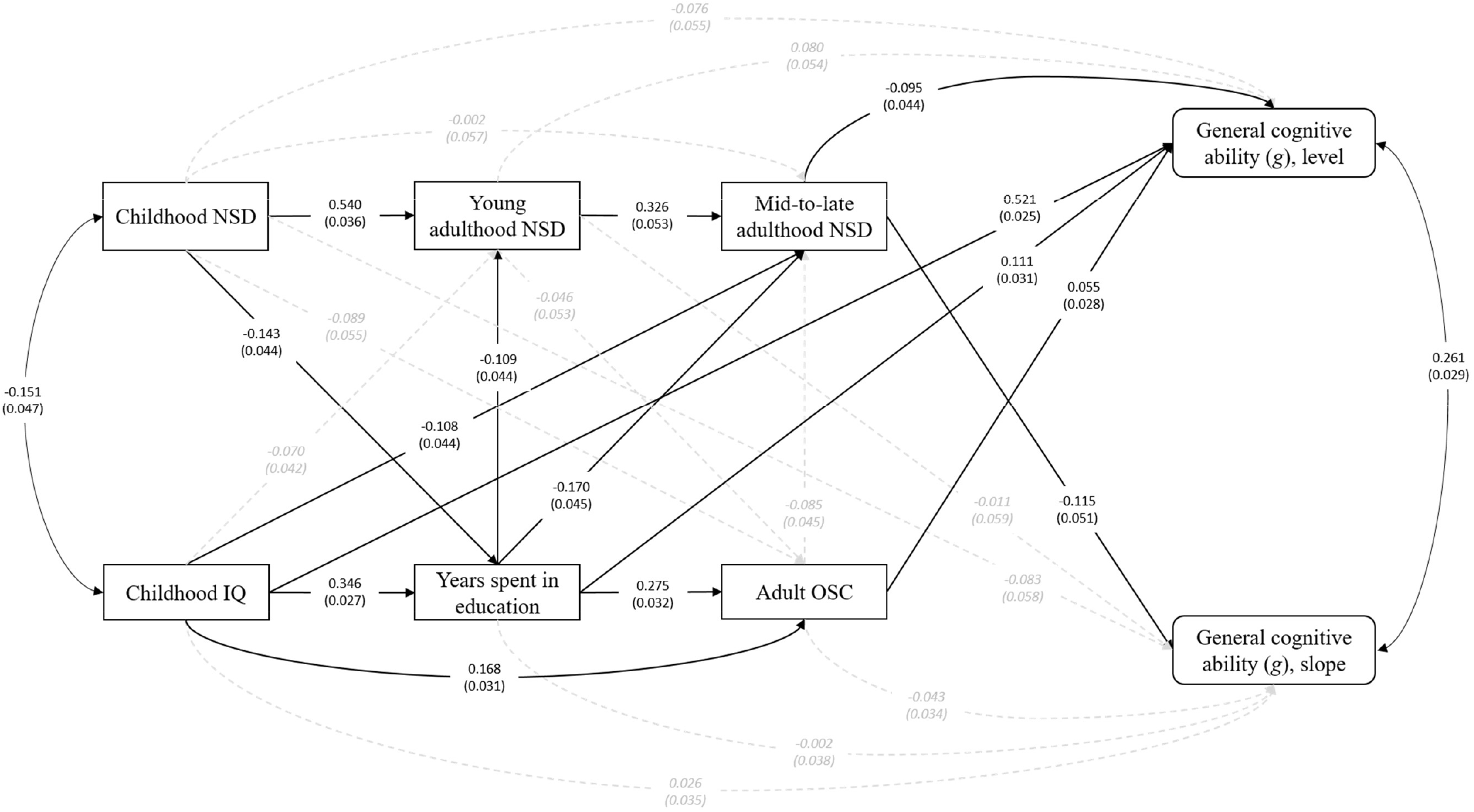
Path diagram depicting life course associations between neighbourhood social deprivation (NSD), general cognitive ability (*g*) and mediators. Black solid lines represents significant (*p*<0.05), grey dashed lines non-significant associations; double headed arrows are correlations. The intercept and slope of *g* was extracted from the ‘factor-of-curves’ measurement model. All presented variables were adjusted for sex, age (time-variant), parental occupational social class, and *APOE* ε4 allele status (see detailed results in Supplementary Table 6).

### Sensitivity analyses

When further adjusting for health-related covariates (i.e. Model 3), the mid-to-late adulthood NSD association was reduced for *g* intercept but remained significant for slopes of *g* (nominally) and processing speed (Supplementary Table 7). Similarly, excluding 56 individuals with cognitive impairment slightly reduced the effect sizes, but key findings remained nominally significant in Model 2 (Supplementary Table 8). Finally, operationalising NSD with residualised scores resulted to the same findings as the main models, with additional nominally significant associations for childhood NSD and slopes of *g* and processing sleep (Supplementary Table 9).

## DISCUSSION

This study examined the relationship between exposure to neighbourhood deprivation across eight decades and level and slope of late-life cognitive function. There are three key findings. First, living in disadvantaged areas in mid-to-late adulthood was associated with lower age 70 level and steeper decline between age 70 and 82 in general cognitive function. Second, all associations initially identified with apparent domain-specific cognitive abilities were due to their shared variance with *g*. Third, childhood area disadvantage was indirectly associated with *g* through education and through later life neighbourhood deprivation.

Living in socially deprived neighbourhoods during the second part of life was associated with lower levels and steeper declines in *g* and processing speed, with point estimates comparable or larger in magnitude than for individual socioeconomic position. Associations were 42% larger in magnitude on the slope, which has been reported earlier.^35^ Research has shown that key features of built and social environment, including local services supporting older and disabled people,36 more bus routes,^36^ greater neighbourhood walking destinations,^37^ more community centres,^35^ better access to parks,^12^ maintained public spaces^35^ and lower levels of social disorder^38^ can slow cognitive decline among older adulthoods. Residing in an advantaged neighbourhood may have additional benefit for cognitive functioning by supporting social participation and informal socialising, and therefore enabling to connection and engagement with others.^8,39^ Since area-level characteristics may further influence mental, physical and social wellbeing and support or inhibit healthy lifestyles, neighbourhood features should be considered as key modifiable predictors of ‘successful’ cognitive ageing.

Partitioning variance between specific and general abilities is a novel contribution to the literature and suggests that associations found between neighbourhood deprivation and specific domains, such as problem solving,^40^ semantic memory,^41^ and processing speed^37^ might be an artefact because of their high correlation with *g*. This is particularly pertinent for processing speed; 100% of the association with mid-to-late adulthood NSD was due to shared variance with *g*.

The path model highlighted that childhood NSD might indirectly contribute to late-life function through individual-level education and selective residential mobility. Neighbourhood poverty has been linked to lower educational attainment among adolescent,^42^ a prominent determinant of late-life cognitive function.^7^ Although the current study operationalised the link between childhood NSD and childhood IQ as a correlation, it is plausible that at least part of the causal direction underpinning this association goes from advantaged childhood neighbourhood to better cognitive performance in schools.^43^ Childhood environment and subsequent educational enrichment may contribute to build up ‘cognitive reserves’ through life, which can support the brain’s active coping and resilience by delaying pathology and age-related decline.^44^ In line with the literature,^45^ we observed significant ‘tracking by area deprivation’ across the life course: childhood environment indirectly contributed to cognitive function through selective residential mobility across the lifespan. From the life-course perspective pathways are consistent with the ‘chain of risk’ hypothesis,^46^ whereby one detrimental exposure (i.e. childhood deprivation) leads to another one (i.e. lower educational attainment/ staying in more deprived areas in young adulthood) and then to another one (i.e. higher neighbourhood deprivation in mid-to-late adulthood), increasing the risk of lower cognitive function and faster decline.

Future research could usefully i) replicate and refine our findings in larger and more diverse cohorts with higher exposure heterogeneity and cognitive measurements from early adulthood onwards; ii) develop a wider range of longitudinal neighbourhood features and examine their contributions to general and domain-specific abilities over the life course; iii) establish causal pathways between neighbourhood exposure and cognition.

### Strengths and limitations

This study benefitted from a comprehensive set of 10 cognitive tests assessed on five occasions over 12 years, residential history covering 8 decades, and valid childhood cognitive scores making it a unique data source to explore life-course associations between neighbourhood and cognitive function. Modelling latent cognitive variables instead of single cognitive test scores provided a more definitive analysis of cognition than previously available, and also reduced the influence of potential measurement error.^34^ Exploring the direct and indirect associations in the path model is a further strength of our study.

There are limitations to acknowledge. First, LBC1936 comprises a self-selected, and relatively healthy and more educated group of individuals. Population-level data suggests that LBC1936 participants had comparatively high childhood IQ^47^ and thus lower risk of mortality.^48^ Second, associations with neighbourhood deprivation were based on 400-500 participants living in Edinburgh—though we used FIML to account for all available data and minimise bias against less healthy participants subject to attrition/incomplete attendance—larger samples would be required to more reliably detect smaller effect sizes relevant to cognitive ageing differences. Third, to provide consistent longitudinal neighbourhood units we used 1961 census ward geographies, which are unlikely to overlap with participants’ self-defined neighbourhoods, resulting in lower precision and potential misclassification. Fourth, health-related covariates were not available prior to age 70 leading to unmeasured confounding. Last, mid-to-late adulthood NSD covers almost 40 years and it is plausible that a more fine-grained temporal analysis would have revealed sensitive periods within this interval; still, we were not able to separate the effects of mid and late adulthood due to their very high correlation.

## Conclusions

To our knowledge, we conducted one of the most comprehensive investigations of the association between life-course neighbourhood deprivation and later-life cognitive function. Findings highlighted that living in less advantaged areas in mid-to-late adulthood was associated with lower level at age 70 and steeper decline in general cognitive ability between age 70 and 82. Future studies should identify neighbourhood features that are pertinent for cognitive ageing differences and establish causal pathways. Given accumulating evidence neighbourhood context can be considered as policy relevant. Supporting successful cognitive ageing starts in childhood by levelling up the gap in education between neighbourhoods, as well as providing and connecting places where physical, social, and mentally stimulating activities could take place.

## Data Availability

The LBCs study data have been the subject of many internal (within the University of Edinburgh) and external collaborations, which are encouraged. Those who have interests in outcomes other than cognitive domains are particularly encouraged to collaborate. Both LBC studies have clear data dictionaries which help researchers to discern whether the variables they wish to use are present; these provide a simple short title for each variable, alongside a longer, common-sense description/provenance of each variable.

https://www.ed.ac.uk/lothian-birth-cohorts

## ACKNOWLEDGEMENTS

The LBC1936 study was conducted according to the Declaration of Helsinki guidelines with ethical permission obtained from the Multi-Centre Research Ethics Committee for Scotland (MREC/01/0/56), Lothian Research Ethics Committee (wave 1, LREC/2003/2/29), and the Scotland A Research Ethics Committee (waves 2-4, 07/MRE00/58). Written consent was obtained from all participants.

The Lothian Birth Cohort 1936 study acknowledges the financial support of NHS Research Scotland (NRS), through Edinburgh Clinical Research Facility. We gratefully acknowledge the contributions of the LBC1936 participants and members of the LBC1936 research team who collect and manage the LBC data.

IJD, NS, SRC and JP obtained and managed the data for the study. GB, IJD, NS, CWT, SRC and JP conceived and designed the study. GB performed the statistical analyses and led the manuscript preparations, drafting and revision. FC supported the data-analyses. All authors participated in the interpretation of the findings, critically revised the manuscript and approved the final version.

This work was supported by the Economic and Social Research Council, UK (ESRC; grant award ES/T003669/1). The LBC1936 study is supported by the Biotechnology and Biological Sciences Research Council (BBSRC) and the ESRC (BB/W008793/1), Age UK (Disconnected Mind project), the US National Institutes of Health (R01AG054628, which supports IJD), and the University of Edinburgh. SRC was supported by a Sir Henry Dale Fellowship jointly funded by the Wellcome Trust and the Royal Society (221890/Z/20/Z).

Data-availability statement: The LBCs’ study data have been the subject of many internal (within the University of Edinburgh) and external collaborations, which are encouraged. Those who have interests in outcomes other than cognitive domains are particularly encouraged to collaborate. Both LBC studies have clear data dictionaries which help researchers to discern whether the variables they wish to use are present; these provide a simple short title for each variable, alongside a longer, common-sense description/provenance of each variable. This information is available on the study website (https://www.ed.ac.uk/lothian-birth-cohorts) alongside comprehensive data grids listing all variables collected throughout both LBC studies and the wave at which they were introduced, an ‘LBC Data Request Form’ and example Data Transfer Agreement. Initially, the Data Request Form is e-mailed to the Lothian Birth Cohorts Director Dr Simon R. Cox for approval (via a panel comprising study co-investigators). Instances where approved projects require transfer of data or materials outside the University of Edinburgh require a formal Data Transfer Agreement or Material Transfer Agreement to be established with the host institution. The process is facilitated by a full-time LBC database manager – there is no charge.

For the purpose of open access, the author has applied a Creative Commons Attribution (CC BY) licence to any Author Accepted Manuscript version arising from this submission.

## Notes

### Competing Interest Statement

The authors have declared no competing interest.

