## Supplementary Material for "Neighbourhood deprivation across eight decades and late-life cognitive function in the Lothian Birth Cohort 1936: A life-course study"

**Supplementary Materials**

**Supplementary Figure 1:** Pearson correlation coefficients between exposure to neighbourhood social deprivation across A) decades and B) epochs

**Supplementary Figure 2:** Directed acyclic graph depicting associations between neighbourhood social deprivation, cognitive abilities and life course covariates

**Supplementary Table 1:** Means and standard deviations of cognitive tests at each wave of testing

**Supplementary Table 2:** Means and standard deviations of cognitive tests at each wave of testing for completers

**Supplementary Table 3:** ‘Factor-of-curves’ models for the association between neighbourhood deprivation and cognitive abilities with covariates

**Supplementary Table 4**: Model fit measures for the hierarchical ‘factor-of-curves’ models

**Supplementary Table 5:** Bifactor model for the association between neighbourhood social deprivation and cognitive abilities

**Supplementary Table 6:** Standardized path coefficients

**Supplementary Table 7:** Factor-of-curves models for the association between neighbourhood social deprivation and cognitive abilities after adjustment for health covariates

**Supplementary Table 8:** Factor-of-curves models for the association between neighbourhood social deprivation and cognitive abilities after excluding individuals with cognitive impairment

**Supplementary Table 9:** Factor-of-curves models for the association between non-residualized neighbourhood social deprivation and cognitive abilities

**Supplementary Figure 1:** Pearson correlation coefficients between exposure to neighbourhood social deprivation across A) decades and B) epochs.

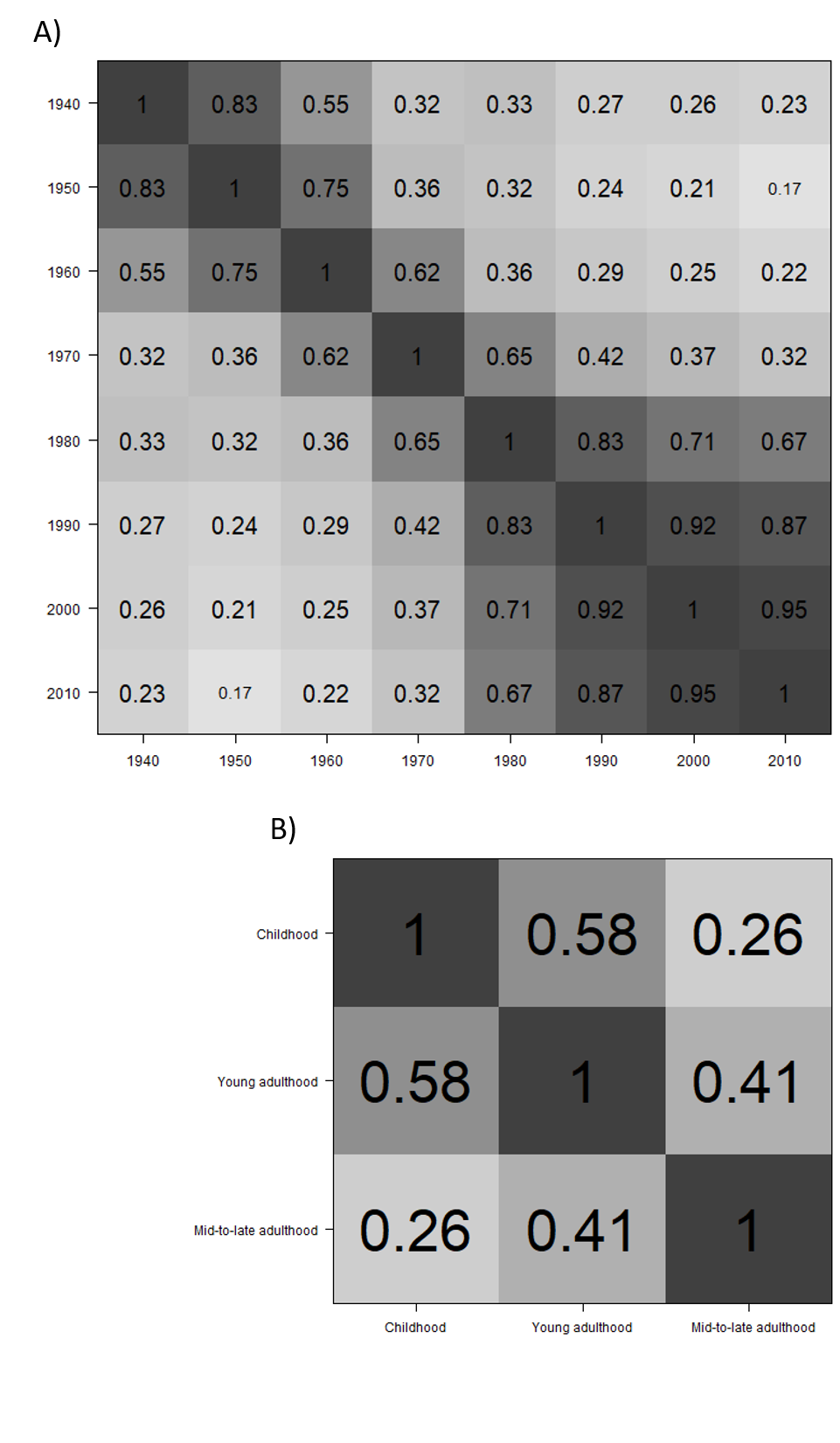

To preserve the maximum amount of available information, pairwise deletion was applied for missing values.

**Supplementary Figure 2:** Directed acyclic graph depicting associations between neighbourhood social deprivation, cognitive abilities and life course covariates. Black arrows present main associations of interest, grey arrows potential confounding pathways; links between covariates are not shown for simplicity. Three sets of covariates were grouped together. Dark grey coloured boxes are likely confounders for all NSD and cognitive function associations (i.e., Model 1 covariates), medium grey coloured covariates present potential pathways on how NSD from early life may be associated with cognitive function (i.e., Model 2 covariates). Light grey coloured health-related covariates may be confounders (as presented in the graph), but could also be on the causal pathway between exposure in later life and outcome, and are therefore included in the sensitivity analysis (i.e., Model 3 covariates). Abbreviations: BMI=body mass index; CVD=cardiovascular diseases; NSD=neighbourhood social deprivation; OSC=occupational social class.

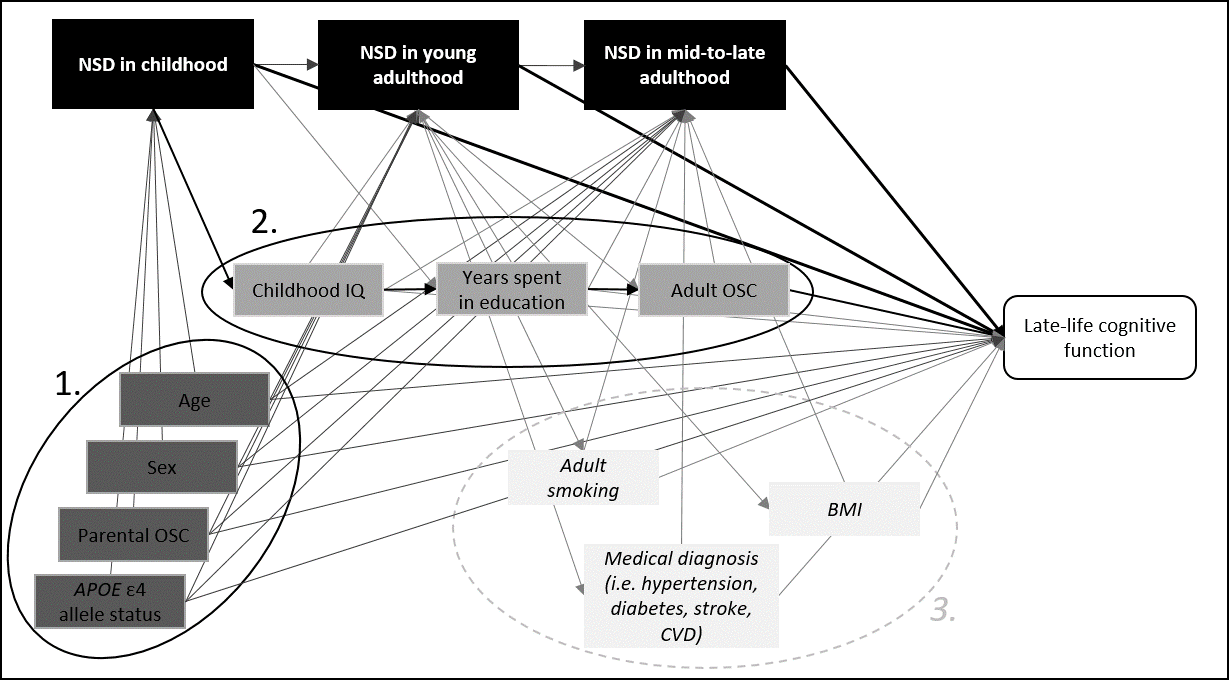

**Supplementary Table 1:** Means and standard deviations of cognitive tests at each wave of testing

| **Cognitive tests** | **Wave 1**  (n=1091) | **Wave 2**  (n=866) | **Wave 3**  (n=697) | **Wave 4**  (n=550) | **Wave 5**  (n=431) |
| --- | --- | --- | --- | --- | --- |
| Visuospatial ability |  |  |  |  |  |
| Matrix Reasoning (mean ± SD) | 13.49 ± 5.13 | 13.17 ± 4.96 | 13.04 ± 4.91 | 12.90 ± 5.03 | 12.93 ± 5.22 |
| Block Design (mean ± SD) | 33.79 ± 10.32 | 33.64 ± 10.08 | 32.18 ± 9.95 | 31.20 ± 9.63 | 29.91 ± 9.60 |
| Spatial Span (mean ± SD) | 14.72 ± 2.83 | 14.69 ± 2.76 | 14.62 ± 2.73 | 14.13 ± 2.72 | 13.89 ± 2.85 |
| Memory |  |  |  |  |  |
| Logical Memory (mean ± SD) | 71.46 ± 17.96 | 74.30 ± 17.88 | 74.58 ± 19.20 | 72.71 ± 20.39 | 72.15 ± 21.52 |
| Verbal Paired Associates (mean ± SD) | 26.44 ± 9.13 | 27.18 ± 9.46 | 26.41 ± 9.56 | 27.14 ± 9.55 | 27.37 ± 9.54 |
| Backward Digit Span (mean ± SD) | 7.73 ± 2.26 | 7.81 ± 2.29 | 7.77 ± 2.37 | 7.56 ± 2.18 | 7.19 ± 2.33 |
| Processing speed |  |  |  |  |  |
| Digit Symbol Substitution (mean ± SD) | 56.60 ± 12.93 | 56.40 ± 12.31 | 53.81 ± 12.93 | 51.24 ± 13.01 | 50.98 ± 12.79 |
| Symbol Search (mean ± SD) | 24.71 ± 6.39 | 24.61 ± 6.18 | 24.60 ± 6.46 | 22.68 ± 6.72 | 22.21 ± 6.93 |
| Four-Choice Reaction Time^a^ (mean ± SD) | 0.64 ± 0.09 | 0.65 ± 0.09 | 0.68 ± 0.10 | 0.71 ± 0.11 | 0.72 ± 0.12 |
| Inspection Time (mean ± SD) | 112.14 ± 11.00 | 111.22 ± 11.79 | 110.14 ± 12.55 | 106.96 ± 13.60 | 106.03 ± 12.72 |

Note: Sample size for specific cognitive tests may vary; therefore, sample size for the respective wave is given. In general, higher test scores indicate better performance.

^a^ Higher values indicate slower reaction time

**Supplementary Table 2:** Means and standard deviations of cognitive tests at each wave of testing for completers (*n*=418)

| **Cognitive tests** | **Wave 1** | **Wave 2** | **Wave 3** | **Wave 4** | **Wave 5** | **Change (wave 1-5)^a^** |
| --- | --- | --- | --- | --- | --- | --- |
| Visuospatial ability |  |  |  |  |  |  |
| Matrix Reasoning (mean ± SD) | 14.77 ± 4.99 | 14.25 ± 4.92 | 13.84 ± 4.85 | 13.44 ± 4.96 | 13.02 ± 5.20 | -1.80 ± 4.52 |
| Block Design (mean ± SD) | 36.06 ± 9.98 | 35.42 ± 10.28 | 33.55 ± 9.89 | 32.52 ± 9.56 | 30.06 ± 9.61 | -6.21 ± 7.56 |
| Spatial Span (mean ± SD) | 15.20 ± 2.79 | 15.06 ± 2.65 | 15.00 ± 2.70 | 14.37 ± 2.77 | 13.95 ± 2.85 | -1.25 ± 2.57 |
| Memory |  |  |  |  |  |  |
| Logical Memory (mean ± SD) | 74.88 ± 17.12 | 77.19 ± 16.79 | 77.18 ± 17.17 | 75.80 ± 18.30 | 72.38 ± 21.55 | -2.69 ± 19.52 |
| Verbal Paired Associates (mean ± SD) | 28.36 ± 8.25 | 28.95 ± 8.83 | 28.16 ± 8.77 | 28.21 ± 9.03 | 27.49 ± 9.53 | -1.25 ± 8.01 |
| Backward Digit Span (mean ± SD) | 8.16 ± 2.35 | 8.18 ± 2.35 | 8.09 ± 2.41 | 7.76 ± 2.19 | 7.22 ± 2.33 | -0.96 ± 1.92 |
| Processing speed |  |  |  |  |  |  |
| Digit Symbol Substitution (mean ± SD) | 60.00 ± 11.98 | 59.70 ± 11.56 | 56.99 ± 11.74 | 53.50 ± 11.99 | 51.08 ± 12.63 | -9.18 ± 9.65 |
| Symbol Search (mean ± SD) | 25.97 ± 6.57 | 25.93 ± 5.79 | 25.87 ± 6.05 | 23.61 ± 6.23 | 22.28 ± 6.90 | -3.87 ± 6.14 |
| Four-Choice Reaction Time^*^ (mean ± SD) | 0.62 ± 0.08 | 0.63 ± 0.08 | 0.66 ± 0.09 | 0.69 ± 0.10 | 0.72 ± 0.12 | 0.10 ± 0.10 |
| Inspection Time (mean ± SD) | 114.06 ± 10.00 | 112.79 ± 11.55 | 111.63 ± 11.56 | 108.79 ± 11.77 | 105.99 ± 12.70 | -8.11 ± 12.20 |

Note: Completers participated in each follow-up wave; sample size for specific cognitive tests may vary. In general, higher test scores indicate better performance.

^a^ Mean and SD of the changes.

^b^ Higher values indicate slower reaction time

**Supplementary Table 3:** ‘Factor-of-curves’ models for the association between neighbourhood deprivation and cognitive abilities with covariates

| **Predictors** | **General cognitive ability (*g*)** | | |  | **Visuospatial ability** | | |  | **Memory** | | |  | **Processing speed** | | |
| --- | --- | --- | --- | --- | --- | --- | --- | --- | --- | --- | --- | --- | --- | --- | --- |
|  | *β* | 95% CI | *p* |  | *β* | 95% CI | *p* |  | *β* | 95% CI | *p* |  | *β* | 95% CI | *p* |
| **Intercept on** |  |  |  |  |  |  |  |  |  |  |  |  |  |  |  |
| Neighbourhood deprivation |  |  |  |  |  |  |  |  |  |  |  |  |  |  |  |
| in childhood | -0.075 | -0.191, 0.040 | 0.203 |  | -0.096 | -0.209, 0.018 | 0.099 |  | -0.032 | -0.168, 0.103 | 0.640 |  | -0.009 | -0.132, 0.114 | 0.888 |
| in young adulthood | 0.087 | -0.027, 0.200 | 0.134 |  | 0.077 | -0.035, 0.188 | 0.177 |  | 0.147 | 0.015, 0.278 | 0.029 |  | 0.005 | -0.117, 0.126 | 0.936 |
| in mid-to-late adulthood | -0.113 | -0.204, -0.021 | 0.016 |  | -0.091 | -0.184, 0.001 | 0.052 |  | -0.094 | -0.204, 0.016 | 0.093 |  | -0.073 | -0.174, 0.029 | 0.160 |
| Sex | -0.105 | -0.160, -0.050 | <0.001 |  | -0.245 | -0.299, -0.191 | <0.001 |  | 0.111 | 0.049, 0.173 | <0.001 |  | 0.014 | -0.045, 0.073 | 0.645 |
| Parental OSC | -0.013 | -0.075, 0.048 | 0.669 |  | -0.040 | -0.103, 0.024 | 0.220 |  | -0.003 | -0.075, 0.070 | 0.939 |  | 0.018 | -0.050, 0.086 | 0.602 |
| *APOE* ε4 allele status | -0.080 | -0.133, -0.027 | 0.003 |  | -0.080 | -0.136, -0.024 | 0.005 |  | -0.033 | -0.096, 0.030 | 0.306 |  | -0.076 | -0.134, -0.018 | 0.010 |
| Childhood IQ | 0.613 | 0.560, 0.665 | <0.001 |  | 0.508 | 0.452, 0.565 | <0.001 |  | 0.578 | 0.512, 0.644 | <0.001 |  | 0.475 | 0.414, 0.535 | <0.001 |
| Years spent in education | 0.128 | 0.063, 0.193 | <0.001 |  | 0.127 | 0.059, 0.195 | <0.001 |  | 0.174 | 0.097, 0.250 | <0.001 |  | 0.054 | -0.018, 0.125 | 0.143 |
| Adult OSC | 0.062 | 0.001, 0.122 | 0.045 |  | 0.076 | 0.013, 0.139 | 0.019 |  | -0.026 | -0.097, 0.046 | 0.483 |  | 0.071 | 0.005, 0.137 | 0.035 |
| **Slope on** |  |  |  |  |  |  |  |  |  |  |  |  |  |  |  |
| Neighbourhood deprivation |  |  |  |  |  |  |  |  |  |  |  |  |  |  |  |
| in childhood | -0.104 | -0.257, 0.048 | 0.180 |  | -0.048 | -0.261, 0.166 | 0.661 |  | -0.044 | -0.194, 0.105 | 0.560 |  | -0.104 | -0.262, 0.053 | 0.195 |
| in young adulthood | 0.007 | -0.147, 0.161 | 0.929 |  | -0.028 | -0.240, 0.184 | 0.795 |  | 0.008 | -0.143, 0.159 | 0.915 |  | 0.016 | -0.143, 0.174 | 0.848 |
| in mid-to-late adulthood | -0.160 | -0.290, -0.031 | 0.015 |  | -0.093 | -0.270, 0.084 | 0.305 |  | 0.018 | -0.114, 0.149 | 0.791 |  | -0.215 | -0.347, -0.083 | 0.001 |
| Sex | 0.105 | 0.024, 0.186 | 0.011 |  | 0.141 | 0.016, 0.265 | 0.026 |  | 0.058 | -0.029, 0.144 | 0.192 |  | 0.102 | 0.015, 0.189 | 0.022 |
| Parental OSC | 0.006 | -0.083, 0.095 | 0.890 |  | 0.095 | -0.038, 0.227 | 0.163 |  | -0.004 | -0.099, 0.091 | 0.932 |  | 0.002 | -0.094, 0.098 | 0.968 |
| *APOE* ε4 allele status | -0.220 | -0.299, -0.141 | <0.001 |  | -0.166 | -0.289, -0.043 | 0.008 |  | -0.238 | -0.323, -0.154 | <0.001 |  | -0.199 | -0.284, -0.115 | <0.001 |
| Childhood IQ | -0.077 | -0.168, 0.015 | 0.100 |  | -0.251 | -0.390, -0.112 | <0.001 |  | -0.019 | -0.117, 0.079 | 0.705 |  | -0.057 | -0.154, 0.041 | 0.254 |
| Years spent in education | -0.027 | -0.123, 0.070 | 0.591 |  | -0.063 | -0.210, 0.085 | 0.405 |  | 0.000 | -0.102, 0.103 | 0.997 |  | -0.024 | -0.128, 0.079 | 0.646 |
| Adult OSC | -0.084 | -0.176, 0.008 | 0.074 |  | -0.069 | -0.211, 0.074 | 0.343 |  | -0.009 | -0.109, 0.091 | 0.863 |  | -0.107 | -0.205, -0.009 | 0.033 |

Models were run for general cognitive ability and for each domain-specific cognitive abilities separately. Effect estimates were expressed as standardized betas (*β*). All models are also adjusted for age (time-variant).

**Supplementary Table 4**: Model fit measures for the ‘factor-of-curves’ models

| **Cognitive domain** | **Model 1** | | | |  | **Model 2** | | | |
| --- | --- | --- | --- | --- | --- | --- | --- | --- | --- |
|  | CFI | TLI | RMSEA | SRMR |  | CFI | TLI | RMSEA | SRMR |
| General cognitive ability (*g*) | 0.938 | 0.934 | 0.030 | 0.059 |  | 0.933 | 0.930 | 0.030 | 0.057 |
| Visuospatial ability | 0.992 | 0.991 | 0.015 | 0.027 |  | 0.989 | 0.987 | 0.017 | 0.027 |
| Memory | 0.953 | 0.947 | 0.036 | 0.033 |  | 0.949 | 0.942 | 0.036 | 0.031 |
| Processing speed | 0.943 | 0.937 | 0.040 | 0.046 |  | 0.940 | 0.934 | 0.038 | 0.045 |

Reference values are: CFI>0.95, TLI>0.95, RMSEA<0.06, SRMR<0.08.^1^ CFI=Comparative Fit Index; TLI= Tucker-Lewis Index; RMSEA=Root Mean Square Error of Approximation; SRMR=Standardized Root Mean Square Residual

Model 1: adjusted for sex, age (time-variant), parental occupational social class, and *APOE* ε4 allele status.

Model 2: adjusted also for childhood IQ, years spent in education, and adult occupational social class.

**Supplementary Table 5:** Bifactor model for the association between neighbourhood social deprivation and cognitive abilities

| Neighbourhood  social deprivation | | Model 1 | | | |  | Model 2 | | | |
| --- | --- | --- | --- | --- | --- | --- | --- | --- | --- | --- |
|  |  | *β* | 95% CI | *p* | *p_FDR_* |  | *β* | 95% CI | *p* | *p_FDR_* |
| **General cognitive ability (*g*)** | | | | | | | | | | |
| Intercept | Childhood | -0.154 | -0.284, -0.024 | 0.021 | 0.168 |  | -0.126 | -0.239, -0.013 | 0.029 | 0.232 |
|  | Young adulthood | 0.046 | -0.087, 0.179 | 0.499 | 0.630 |  | 0.050 | -0.066, 0.165 | 0.399 | 0.871 |
|  | Mid-to-late adulthood | **-0.197** | **-0.300, -0.093** | **<0.001** | **0.007** |  | -0.142 | -0.237, -0.046 | 0.004 | 0.096 |
| Slope | Childhood | -0.080 | -0.233, 0.073 | 0.306 | 0.589 |  | -0.096 | -0.246, 0.053 | 0.206 | 0.600 |
|  | Young adulthood | 0.042 | -0.113, 0.196 | 0.597 | 0.716 |  | 0.011 | -0.141, 0.162 | 0.892 | 0.992 |
|  | Mid-to-late adulthood | -0.109 | -0.233, 0.016 | 0.088 | 0.466 |  | -0.160 | -0.284, -0.035 | 0.012 | 0.144 |
| **Visuospatial ability** | | | | | | | | | | |
| Intercept | Childhood | 0.081 | -0.088, 0.251 | 0.345 | 0.589 |  | 0.115 | -0.049, 0.279 | 0.168 | 0.576 |
|  | Young adulthood | -0.069 | -0.236, 0.098 | 0.419 | 0.589 |  | -0.041 | -0.204, 0.123 | 0.627 | 0.895 |
|  | Mid-to-late adulthood | -0.089 | -0.225, 0.046 | 0.196 | 0.534 |  | -0.022 | -0.161, 0.117 | 0.758 | 0.957 |
| Slope | Childhood | 0.120 | -0.160, 0.401 | 0.400 | 0.589 |  | 0.084 | -0.177, 0.346 | 0.527 | 0.895 |
|  | Young adulthood | 0.031 | -0.254, 0.316 | 0.832 | 0.880 |  | 0.003 | -0.261, 0.268 | 0.980 | 0.992 |
|  | Mid-to-late adulthood | 0.136 | -0.090, 0.362 | 0.238 | 0.534 |  | 0.069 | -0.151, 0.288 | 0.540 | 0.895 |
| **Memory** | | | | | | | | | | |
| Intercept | Childhood | 0.065 | -0.101, 0.231 | 0.442 | 0.589 |  | 0.074 | -0.075, 0.223 | 0.330 | 0.792 |
|  | Young adulthood | 0.024 | -0.140, 0.188 | 0.775 | 0.880 |  | 0.092 | -0.057, 0.242 | 0.225 | 0.600 |
|  | Mid-to-late adulthood | -0.153 | -0.279, -0.026 | 0.018 | 0.168 |  | -0.030 | -0.155, 0.095 | 0.634 | 0.895 |
| Slope | Childhood | 0.016 | -0.170, 0.201 | 0.868 | 0.880 |  | 0.020 | -0.162, 0.203 | 0.827 | 0.992 |
|  | Young adulthood | -0.014 | -0.199, 0.171 | 0.880 | 0.880 |  | -0.010 | -0.192, 0.173 | 0.917 | 0.992 |
|  | Mid-to-late adulthood | 0.117 | -0.030, 0.263 | 0.120 | 0.480 |  | 0.134 | -0.016, 0.284 | 0.080 | 0.320 |
| **Processing speed** | | | | | | | | | | |
| Intercept | Childhood | 0.119 | -0.047, 0.284 | 0.160 | 0.534 |  | 0.140 | -0.016, 0.296 | 0.079 | 0.320 |
|  | Young adulthood | -0.099 | -0.265, 0.068 | 0.245 | 0.534 |  | -0.029 | -0.189, 0.130 | 0.717 | 0.956 |
|  | Mid-to-late adulthood | -0.082 | -0.215, 0.051 | 0.228 | 0.534 |  | 0.039 | -0.093, 0.172 | 0.562 | 0.895 |
| Slope | Childhood | -0.127 | -0.381, 0.128 | 0.330 | 0.589 |  | -0.092 | -0.340, 0.157 | 0.470 | 0.895 |
|  | Young adulthood | 0.209 | -0.038, 0.456 | 0.097 | 0.467 |  | 0.233 | -0.005, 0.471 | 0.055 | 0.320 |
|  | Mid-to-late adulthood | -0.093 | -0.297, 0.112 | 0.376 | 0.589 |  | 0.001 | -0.206, 0.208 | 0.992 | 0.992 |

*Note*: Bifactor models estimated the associations for general ability and domain-specific cognitive abilities simultaneously. Effect estimates were expressed as standardized betas (*β*). Bold typeface denotes FDR-corrected significance.

Model 1: adjusted for sex, age (time-variant), parental occupational social class, and *APOE* ε4 allele status.

Model 2: adjusted also for childhood IQ, years spent in education, and adult occupational social class.

**Supplementary Table 6:** Standardized path coefficients

| **Associations** | **Path** |  | ***β*** | **SE** | ***p*** |
| --- | --- | --- | --- | --- | --- |
| G intercept | ~ | Childhood NSD | -0.076 | 0.055 | 0.170 |
| G intercept | ~ | Young adulthood NSD | 0.080 | 0.054 | 0.137 |
| **G intercept** | **~** | **Mid-to-late adulthood NSD** | **-0.095** | **0.044** | **0.031** |
| **G intercept** | **~** | **Childhood IQ** | **0.521** | **0.025** | **0.000** |
| **G intercept** | **~** | **Years spent in education** | **0.111** | **0.031** | **0.000** |
| **G intercept** | **~** | **Adult OSC** | **0.055** | **0.028** | **0.047** |
| G intercept | ~ | Parental OSC | -0.015 | 0.028 | 0.593 |
| **G intercept** | **~** | **Sex** | **-0.098** | **0.024** | **0.000** |
| **G intercept** | **~** | ***APOE* ε4 allele status** | **-0.090** | **0.025** | **0.000** |
| G slope | ~ | Childhood NSD | -0.083 | 0.058 | 0.154 |
| G slope | ~ | Young adulthood NSD | 0.011 | 0.059 | 0.858 |
| **G slope** | **~** | **Mid-to-late adulthood NSD** | **-0.115** | **0.051** | **0.023** |
| G slope | ~ | Childhood IQ | 0.026 | 0.035 | 0.451 |
| G slope | ~ | Years spent in education | -0.002 | 0.038 | 0.451 |
| G slope | ~ | Adult OSC | -0.043 | 0.034 | 0.214 |
| G slope | ~ | Parental OSC | -0.002 | 0.034 | 0.953 |
| **G slope** | **~** | **Sex** | **0.061** | **0.030** | **0.043** |
| **G slope** | **~** | ***APOE* ε4 allele status** | **-0.170** | **0.030** | **0.000** |
| **G slope** | **~~** | **G intercept** | **0.261** | **0.029** | **0.000** |
| Mid-to-late adulthood NSD | ~ | Childhood NSD | -0.002 | 0.057 | 0.978 |
| **Mid-to-late adulthood NSD** | **~** | **Young adulthood NSD** | **0.326** | **0.053** | **0.000** |
| **Mid-to-late adulthood NSD** | **~** | **Childhood IQ** | **-0.108** | **0.044** | **0.015** |
| **Mid-to-late adulthood NSD** | **~** | **Years spent in education** | **-0.170** | **0.045** | **0.000** |
| Mid-to-late adulthood NSD | ~ | Adult OSC | -0.085 | 0.045 | 0.055 |
| Mid-to-late adulthood NSD | ~ | Parental OSC | -0.025 | 0.043 | 0.562 |
| **Mid-to-late adulthood NSD** | **~** | **Sex** | **0.079** | **0.039** | **0.044** |
| Mid-to-late adulthood NSD | ~ | *APOE* ε4 allele status | 0.036 | 0.040 | 0.377 |
| **Young adulthood NSD** | **~** | **Childhood NSD** | **0.540** | **0.036** | **0.000** |
| Young adulthood NSD | ~ | Childhood IQ | -0.070 | 0.042 | 0.097 |
| **Young adulthood NSD** | **~** | **Years spent in education** | **-0.109** | **0.044** | **0.013** |
| Young adulthood NSD | ~ | Parental OSC | -0.044 | 0.041 | 0.281 |
| Young adulthood NSD | ~ | Sex | 0.005 | 0.037 | 0.901 |
| Young adulthood NSD | ~ | *APOE* ε4 allele status | 0.013 | 0.039 | 0.746 |
| Adult OSC | ~ | Childhood NSD | -0.089 | 0.055 | 0.110 |
| Adult OSC | ~ | Young adulthood NSD | -0.046 | 0.053 | 0.379 |
| **Adult OSC** | **~** | **Childhood IQ** | **0.168** | **0.031** | **0.000** |
| **Adult OSC** | **~** | **Years spent in education** | **0.275** | **0.032** | **0.000** |
| **Adult OSC** | **~** | **Parental OSC** | **0.078** | **0.032** | **0.014** |
| Adult OSC | ~ | Sex | 0.046 | 0.027 | 0.094 |
| Adult OSC | ~ | *APOE* ε4 allele status | 0.031 | 0.028 | 0.273 |
| **Years spent in education** | **~** | **Childhood NSD** | **-0.143** | **0.044** | **0.001** |
| **Years spent in education** | **~** | **Childhood IQ** | **0.346** | **0.027** | **0.000** |
| **Years spent in education** | **~** | **Parental OSC** | **0.244** | **0.029** | **0.000** |
| Years spent in education | ~ | Sex | -0.027 | 0.027 | 0.320 |
| Years spent in education | ~ | *APOE* ε4 allele status | 0.021 | 0.027 | 0.438 |
| **Childhood IQ** | **~** | **Parental OSC** | **0.221** | **0.031** | **0.000** |
| **Childhood IQ** | **~** | **Sex** | **0.069** | **0.030** | **0.021** |
| Childhood IQ | ~ | *APOE* ε4 allele status | 0.016 | 0.031 | 0.594 |
| **Childhood NSD** | **~** | **Parental OSC** | **-0.220** | **0.046** | **0.000** |
| Childhood NSD | ~ | Sex | -0.039 | 0.047 | 0.396 |
| Childhood NSD | ~ | *APOE* ε4 allele status | 0.049 | 0.049 | 0.319 |
| **Childhood IQ** | **~~** | **Childhood NSD** | **-0.151** | **0.047** | **0.001** |
| G intercept | ~~ | G intercept | 0.588 | 0.024 | 0.000 |
| G slope | ~~ | G slope | 0.940 | 0.016 | 0.000 |
| Mid-to-late adulthood NSD | ~~ | Mid-to-late adulthood NSD | 0.736 | 0.035 | 0.000 |
| Young adulthood NSD | ~~ | Young adulthood NSD | 0.622 | 0.038 | 0.000 |
| Childhood NSD | ~~ | Childhood NSD | 0.947 | 0.021 | 0.000 |
| Adult OSC | ~~ | Adult OSC | 0.778 | 0.024 | 0.000 |
| Years spent in education | ~~ | Years spent in education | 0.730 | 0.025 | 0.000 |
| Childhood IQ | ~~ | Childhood IQ | 0.948 | 0.014 | 0.000 |
| Parental OSC | ~~ | Parental OSC | 1.000 | 0.000 | NA |
| Parental OSC | ~~ | Sex | -0.030 | 0.000 | NA |
| Parental OSC | ~~ | *APOE* ε4 allele status | -0.048 | 0.000 | NA |
| Sex | ~~ | Sex | 1.000 | 0.000 | NA |
| Sex | ~~ | *APOE* ε4 allele status | -0.040 | 0.000 | NA |
| *APOE* ε4 allele status | ~~ | *APOE* ε4 allele status | 1.000 | 0.000 | NA |
| G intercept | ~ | 1 | 6.056 | 0.198 | 0.000 |
| G slope | ~ | 1 | -2.135 | 0.202 | 0.000 |
| Mid-to-late adulthood NSD | ~ | 1 | -0.597 | 0.244 | 0.014 |
| Young adulthood NSD | ~ | 1 | -0.261 | 0.205 | 0.203 |
| Childhood NSD | ~ | 1 | 0.691 | 0.249 | 0.006 |
| Adult OSC | ~ | 1 | 2.670 | 0.171 | 0.000 |
| Years spent in education | ~ | 1 | -0.661 | 0.148 | 0.000 |
| Childhood IQ | ~ | 1 | -0.884 | 0.162 | 0.000 |
| Parental OSC | ~ | 1 | 2.844 | 0.000 | NA |
| Sex | ~ | 1 | 2.995 | 0.000 | NA |
| *APOE* ε4 allele status | ~ | 1 | 2.837 | 0.000 | NA |

**Supplementary Table 7**: ‘Factor-of-curves’ models for the association between neighbourhood social deprivation and cognitive abilities after adjustment for health covariates

| Neighbourhood  social deprivation | | **Model 3** | | | |
| --- | --- | --- | --- | --- | --- |
|  |  | *β* | 95% CI | *p* | *p_FDR_* |
| **General cognitive ability (*g*)** | | | | | |
| Intercept | Childhood | -0.071 | -0.186, 0.044 | 0.227 | 0.272 |
|  | Young adulthood | 0.075 | -0.037, 0.187 | 0.190 | 0.272 |
|  | Mid-to-late adulthood | -0.090 | -0.181, 0.002 | 0.056 | 0.168 |
| Slope | Childhood | -0.097 | -0.251, 0.056 | 0.214 | 0.272 |
|  | Young adulthood | 0.011 | -0.144, 0.167 | 0.885 | 0.885 |
|  | Mid-to-late adulthood | -0.155 | -0.287, -0.023 | 0.021 | 0.126 |
| **Visuospatial ability** | | | | | |
| Intercept | Childhood | -0.086 | -0.199, 0.028 | 0.138 | 0.497 |
|  | Young adulthood | 0.059 | -0.052, 0.170 | 0.298 | 0.766 |
|  | Mid-to-late adulthood | -0.077 | -0.170, 0.016 | 0.105 | 0.473 |
| Slope | Childhood | -0.043 | -0.258, 0.172 | 0.697 | 0.985 |
|  | Young adulthood | -0.007 | -0.222, 0.209 | 0.953 | 0.985 |
|  | Mid-to-late adulthood | -0.076 | -0.258, 0.105 | 0.412 | 0.824 |
| **Memory** | | | | | |
| Intercept | Childhood | -0.042 | -0.178, 0.094 | 0.545 | 0.981 |
|  | Young adulthood | 0.151 | 0.018, 0.283 | 0.026 | 0.234 |
|  | Mid-to-late adulthood | -0.095 | -0.207, 0.016 | 0.095 | 0.473 |
| Slope | Childhood | -0.039 | -0.189, 0.111 | 0.612 | 0.985 |
|  | Young adulthood | -0.005 | -0.157, 0.147 | 0.951 | 0.985 |
|  | Mid-to-late adulthood | 0.021 | -0.113, 0.154 | 0.762 | 0.985 |
| **Processing speed** | | | | | |
| Intercept | Childhood | -0.006 | -0.127, 0.114 | 0.919 | 0.985 |
|  | Young adulthood | -0.001 | -0.120, 0.118 | 0.985 | 0.985 |
|  | Mid-to-late adulthood | -0.044 | -0.144, 0.057 | 0.392 | 0.824 |
| Slope | Childhood | -0.097 | -0.255, 0.061 | 0.227 | 0.681 |
|  | Young adulthood | 0.022 | -0.137, 0.181 | 0.787 | 0.985 |
|  | Mid-to-late adulthood | **-0.211** | **-0.345, -0.077** | **0.002** | **0.036** |

*Note*: Models were run for general ability and for each domain-specific cognitive abilities separately. Effect estimates were expressed as standardized betas (*β*). Bold typeface denotes FDR-corrected significance.

Model 3: adjusted for sex, age (time-variant), parental occupational social class, *APOE* ε4 allele status, childhood IQ, years spent in education, adult occupational social class, smoking status at age 70, BMI at age 70, and history of hypertension, diabetes, stroke and cardiovascular diseases.

**Supplementary Table 8:** ‘Factor-of-curves’ models for the association between neighbourhood social deprivation and cognitive abilities after excluding individuals with cognitive impairment (*n*=1035)

| Neighbourhood  social deprivation | | **Model 1** | | | |  | **Model 2** | | | |
| --- | --- | --- | --- | --- | --- | --- | --- | --- | --- | --- |
|  |  | *β* | 95% CI | *p* | *p_FDR_* |  | *β* | 95% CI | *p* | *p_FDR_* |
| **General cognitive ability (*g*)** | | | | | | | | | | |
| Level | Childhood | -0.140 | -0.267, -0.013 | 0.031 | 0.093 |  | -0.078 | -0.194, 0.037 | 0.183 | 0.220 |
|  | Young adulthood | 0.015 | -0.113, 0.143 | 0.814 | 0.814 |  | 0.083 | -0.031, 0.196 | 0.152 | 0.220 |
|  | Mid-to-late adulthood | **-0.275** | **-0.378, -0.173** | **<0.001** | **<0.001** |  | -0.112 | -0.207, -0.018 | 0.020 | 0.075 |
| Slope | Childhood | -0.107 | -0.265, 0.052 | 0.186 | 0.279 |  | -0.118 | -0.276, 0.041 | 0.145 | 0.220 |
|  | Young adulthood | 0.055 | -0.104, 0.215 | 0.497 | 0.596 |  | 0.042 | -0.116, 0.200 | 0.605 | 0.605 |
|  | Mid-to-late adulthood | -0.124 | -0.255, 0.006 | 0.062 | 0.124 |  | -0.153 | -0.286, -0.019 | 0.025 | 0.075 |
| **Visuospatial ability** | | | | | | | | | | |
| Level | Childhood | -0.146 | -0.274, -0.018 | 0.026 | 0.094 |  | -0.097 | -0.211, 0.016 | 0.092 | 0.414 |
|  | Young adulthood | 0.016 | -0.113, 0.144 | 0.812 | 0.937 |  | 0.068 | -0.043, 0.180 | 0.230 | 0.520 |
|  | Mid-to-late adulthood | **-0.243** | **-0.346, -0.140** | **<0.001** | **<0.001** |  | -0.096 | -0.190, -0.001 | 0.047 | 0.282 |
| Slope | Childhood | 0.009 | -0.240, 0.257 | 0.947 | 0.948 |  | -0.006 | -0.247, 0.235 | 0.960 | 0.960 |
|  | Young adulthood | 0.057 | -0.190, 0.303 | 0.651 | 0.901 |  | 0.034 | -0.205, 0.273 | 0.780 | 0.936 |
|  | Mid-to-late adulthood | -0.006 | -0.200, 0.187 | 0.948 | 0.948 |  | -0.088 | -0.282, 0.106 | 0.375 | 0.750 |
| **Memory** | | | | | | | | | | |
| Level | Childhood | -0.070 | -0.223, 0.082 | 0.364 | 0.728 |  | -0.029 | -0.166, 0.109 | 0.684 | 0.936 |
|  | Young adulthood | 0.078 | -0.073, 0.230 | 0.313 | 0.704 |  | 0.136 | 0.002, 0.270 | 0.046 | 0.282 |
|  | Mid-to-late adulthood | **-0.223** | **-0.345, -0.101** | **<0.001** | **0.003** |  | -0.070 | -0.185, 0.045 | 0.231 | 0.520 |
| Slope | Childhood | -0.021 | -0.175, 0.134 | 0.794 | 0.937 |  | -0.016 | -0.171, 0.139 | 0.839 | 0.943 |
|  | Young adulthood | 0.017 | -0.140, 0.173 | 0.833 | 0.937 |  | 0.030 | -0.126, 0.185 | 0.707 | 0.936 |
|  | Mid-to-late adulthood | 0.039 | -0.093, 0.171 | 0.563 | 0.876 |  | 0.051 | -0.086, 0.187 | 0.468 | 0.842 |
| **Processing speed** | | | | | | | | | | |
| Level | Childhood | -0.074 | -0.207, 0.059 | 0.277 | 0.704 |  | -0.020 | -0.143, 0.102 | 0.745 | 0.936 |
|  | Young adulthood | -0.037 | -0.172, 0.097 | 0.584 | 0.876 |  | 0.005 | -0.117, 0.127 | 0.940 | 0.960 |
|  | Mid-to-late adulthood | **-0.203** | **-0.312, -0.094** | **<0.001** | **0.003** |  | -0.078 | -0.183, 0.027 | 0.144 | 0.432 |
| Slope | Childhood | -0.118 | -0.283, 0.047 | 0.161 | 0.483 |  | -0.127 | -0.291, 0.036 | 0.127 | 0.432 |
|  | Young adulthood | 0.054 | -0.113, 0.220 | 0.528 | 0.876 |  | 0.036 | -0.129, 0.201 | 0.669 | 0.936 |
|  | Mid-to-late adulthood | **-0.177** | **-0.313, -0.041** | **0.011** | **0.049** |  | -0.206 | -0.345, -0.067 | 0.004 | 0.072 |

*Note*: Cognitive impairment was defined as either having a diagnosis of dementia or scoring <24 points in the Mini Mental State Examination in any of the five included survey waves (n=56). Models were run for general ability and for each domain-specific cognitive abilities separately. Effect estimates were expressed as standardized betas (*β*). Bold typeface denotes FDR-corrected significance.

Model 1: adjusted for sex, age (time-variant), parental occupational social class, and *APOE* ε4 allele status.

Model 2: adjusted also for childhood IQ, years spent in education, and adult occupational social class.

**Supplementary Table 9:** ‘Factor-of-curves’ models for the association between residualized neighbourhood social deprivation and cognitive abilities

| Neighbourhood  social deprivation | | **Model 1** | | | |  | **Model 2** | | | |
| --- | --- | --- | --- | --- | --- | --- | --- | --- | --- | --- |
|  |  | *β* | 95% CI | *p* | *p_FDR_* |  | *β* | 95% CI | *p* | *p_FDR_* |
| **General cognitive ability (*g*)** | | | | | | | | | | |
| Intercept | Childhood | **-0.171** | **-0.278, -0.064** | **0.002** | **0.006** |  | -0.027 | -0.125, 0.071 | 0.585 | 0.585 |
|  | Young adulthood | -0.061 | 0.168, 0.046 | 0.265 | 0.318 |  | 0.064 | -0.032, 0.159 | 0.190 | 0.285 |
|  | Mid-to-late adulthood | **-0.266** | **-0.361, -0.171** | **<0.001** | **<0.001** |  | -0.104 | -0.192, -0.017 | 0.020 | 0.060 |
| Slope | Childhood | -0.098 | -0.222, 0.027 | 0.123 | 0.185 |  | -0.138 | -0.266, -0.010 | 0.034 | 0.068 |
|  | Young adulthood | -0.014 | -0.138, 0.110 | 0.824 | 0.824 |  | -0.048 | -0.174, 0.079 | 0.458 | 0.550 |
|  | Mid-to-late adulthood | -0.118 | -0.237, 0.000 | 0.050 | 0.100 |  | -0.162 | -0.284, -0.039 | 0.010 | 0.060 |
| **Visuospatial ability** | | | | | | | | | | |
| Intercept | Childhood | -0.160 | -0.266, -0.053 | 0.003 | 0.014 |  | -0.042 | -0.139, 0.055 | 0.397 | 0.596 |
|  | Young adulthood | -0.031 | -0.137, 0.074 | 0.559 | 0.857 |  | 0.073 | -0.021, 0.167 | 0.126 | 0.378 |
|  | Mid-to-late adulthood | **-0.233** | **-0.328, -0.138** | **<0.001** | **<0.001** |  | -0.085 | -0.173, 0.003 | 0.057 | 0.257 |
| Slope | Childhood | -0.054 | -0.235, 0.126 | 0.554 | 0.857 |  | -0.107 | -0.289, 0.075 | 0.250 | 0.563 |
|  | Young adulthood | -0.032 | -0.209, 0.145 | 0.721 | 0.928 |  | -0.083 | -0.261, 0.094 | 0.358 | 0.596 |
|  | Mid-to-late adulthood | -0.016 | -0.185, 0.153 | 0.852 | 0.959 |  | -0.094 | -0.266, 0.079 | 0.286 | 0.572 |
| **Memory** | | | | | | | | | | |
| Intercept | Childhood | -0.060 | -0.185, 0.065 | 0.350 | 0.700 |  | 0.051 | -0.064, 0.167 | 0.385 | 0.596 |
|  | Young adulthood | 0.003 | -0.120, 0.125 | 0.965 | 0.965 |  | 0.110 | -0.001, 0.221 | 0.052 | 0.257 |
|  | Mid-to-late adulthood | **-0.220** | **-0.331, -0.109** | **<0.001** | **0.001** |  | -0.077 | -0.183, 0.029 | 0.153 | 0.393 |
| Slope | Childhood | -0.036 | -0.158, 0.087 | 0.571 | 0.857 |  | -0.037 | -0.164, 0.091 | 0.574 | 0.646 |
|  | Young adulthood | 0.006 | -0.115, 0.127 | 0.919 | 0.965 |  | 0.010 | -0.114, 0.134 | 0.873 | 0.873 |
|  | Mid-to-late adulthood | -0.022 | -0.141, 0.098 | 0.722 | 0.928 |  | -0.017 | -0.143, 0.108 | 0.786 | 0.832 |
| **Processing speed** | | | | | | | | | | |
| Intercept | Childhood | -0.135 | -0.243, -0.027 | 0.014 | 0.042 |  | -0.032 | -0.136, 0.072 | 0.550 | 0.646 |
|  | Young adulthood | -0.110 | -0.216, -0.003 | 0.043 | 0.111 |  | -0.032 | -0.133, 0.068 | 0.529 | 0.646 |
|  | Mid-to-late adulthood | **-0.191** | **-0.289, -0.093** | **<0.001** | **0.001** |  | -0.077 | -0.172, 0.018 | 0.113 | 0.378 |
| Slope | Childhood | -0.101 | -0.230, 0.029 | 0.128 | 0.288 |  | -0.140 | -0.273, -0.007 | 0.039 | 0.257 |
|  | Young adulthood | -0.013 | -0.141, 0.115 | 0.837 | 0.959 |  | -0.047 | -0.177, 0.083 | 0.478 | 0.646 |
|  | Mid-to-late adulthood | **-0.170** | **-0.290, -0.050** | **0.005** | **0.018** |  | **-0.204** | **-0.328, -0.081** | **0.001** | **0.018** |

*Note*: Models were run for general ability and for each domain-specific cognitive abilities separately. Effect estimates were expressed as standardized betas (*β*). Bold typeface denotes FDR-corrected significance.

Model 1: adjusted for sex, age (time-variant), parental occupational social class, and *APOE* ε4 allele status.

Model 2: adjusted also for childhood IQ, years spent in education, and adult occupational social class.
